# Rural trauma team development amongst medical trainees and traffic law enforcement professionals in a low-income country: A protocol for a prospective multi-center interrupted time series of interventional training

**DOI:** 10.1101/2023.09.01.23294946

**Authors:** Herman Lule, Michael Mugerwa, Robinson SSebuufu, Patrick Kyamanywa, Jussi P. Posti, Michael Lowery Wilson

## Abstract

**Background:** Road traffic injuries and their resulting mortality disproportionately affect rural communities in low-middle-income countries (LMICs) due to limited human and infrastructural resources for post-crash care. Evidence from high income countries show that trauma team development training could improve the efficiency, care, and outcome of injuries. A paucity of studies has evaluated the feasibility and applicability of this concept in resource constrained settings. The aim of this study protocol is to establish the feasibility of rural trauma team development and training in a cohort of medical trainees and traffic law enforcement professionals in Uganda.

**Methods:** Muti-center interrupted time series of prospective interventional trainings, using the rural trauma team development course (RTTDC) model of the American College of Surgeons. A team of surgeon consultants will execute the training. A prospective cohort of participants will complete a before and after training validated trauma related multiple choice questionnaire (MCQ) during September 2019-November 2023. The difference in mean pre-post training percentage MCQ scores will be compared using ANOVA-test at 95% CI. Time series regression models will be used to test for autocorrelations in performance. Acceptability and relevance of the training will be assessed using 3- and 5-point Likert scales, respectively. All analyses will be performed using Stata 15.0. Ethical approval was obtained from Research and Ethics Committee of Mbarara University of Science and Technology (Ref: MUREC 1/7, 05/05-19) and Uganda National Council for Science and Technology (Ref: SS 5082) prior recruitment. Retrospective registration was accomplished with Research Registry (UIN: researchregistry9490).

**Highlights:**

- Injury related mortality significantly impact LMICs.
- Rural trauma teams could improve the efficiency of injury care in LMICs.
- This study will assess the effect of rural trauma team development training, viewing medical trainees and traffic police as potential human resources for health.
- The results could inform the implementation of future trauma teams in rural settings.

## Introduction

Trauma is a public health threat which claims an annual death toll of 4.5 million lives globally, contributing to 10% of global disease burden ^1^. Over 90% of global deaths due to trauma disproportionately occur in low- and middle-income countries (LMICs) such as those in sub-Saharan Africa where trauma life support educational programs are scarce ^1^. The main cause of trauma related deaths in Africa are road traffic crashes ^2^. Notwithstanding injury prevention being on top of the agenda for the African Centers for Disease Control, Africa records the highest road traffic crash fatality rates (26.6 per 100,000 population) amongst the world health organization (WHO) regions, which is triple that of Europe at 9.3 per 100,000 ^3^. According to Lagarde et al^4^, sub-Saharan Africa contributes to 10% of the global burden of road traffic crash fatalities despite disproportionately owning only 4% of the world’s motorized vehicles.

Several factors contribute to higher post-crash fatalities in LMICs, including limited: infrastructural, financial, physical, and human resources ^5^. According to the recent WHO (2023) alert on safeguards for health worker recruitment ^5^, it turned out that 67.3% (37 of 55) vulnerable countries for shortage of health workers were from the African region, implying Africa’s obvious slow progress to achieving the sustainable development goal (SDG) target for universal health care coverage by 2030. For instance, a study that evaluated surgical capacity in three African countries found that nurses and medical assistants performed surgical procedures as opposed to surgeons ^6^. However, such “task-shifting” without capacity building through skills-training could have a negative impact on treatment outcomes. Moreover, according to the WHO Fifth Global Forum on Human Resources for Health held in (2023), the deficit of skilled workforce in Africa is anticipated to be exacerbated by the accelerated exodus of health professionals due to Covid-19 related international recruitment, further crippling the fragile health systems ^5^.

### Rationale for rural trauma team development and training in Uganda

Uganda is one of the countries whose universal health care coverage index is below 55 and is on WHO, 2023 safeguard list ^5^. Furthermore, Uganda has no formal pre-hospital care systems thus most patients who sustain trauma arrive at hospitals by public means or by police cars without receiving any primary care which exacerbates their injury severity ^7^. In addition, the country has a health workforce to population density ratio of 1: 25,000 far below the global median of 49 per 10,000 people ^5^. For instance, a study of 72 hospitals in western Uganda revealed that there were only 0.7 surgeons for every 100,000 population and that mostly anesthesia was provided by non-physician anesthetists ^8^. As such, barely any of the Ugandan assessed hospitals met the minimum WHO staffing standards for essential surgery ^9^.

Evidence from systematic reviews show that the lack of sustainable expertise for injury care and inadequate skill-based training of first responders contributes to higher injury-related mortality in LMICs ^10^. Reliably trained and motivated staff are key elements of effective trauma systems ^11^, but are inadequate in LMICs ^12^. There have been efforts from WHO to design a mentorship programme for violence and injury prevention (Mentor-VIP) to address the human resource capacity for health in LMICs ^13^, but this target qualified public health practitioners and established researchers as opposed to clinicians in practice and trainees such as medical interns who make the first contact with injured patients in LMICs. Moreover, task shifting was identified as one of the options to boost human resource workforce in the health sector of LMICs ^14^ but the potential role of medical trainees and traffic police as “sustainable human resources for health in the trauma context” has been underexplored. Several trauma care educational modules have been piloted in LMICs to address the unmet human resource needs but their impact assessment has been theoretical without the skills component ^15^. Moreover, other than the ongoing clinical trial on the effect of advanced life support and primary trauma courses in India ^1^, such trauma education programs have not been validated in randomized controlled settings, which could have affected their clinical application ^16^.

Uganda has a six-year long undergraduate medical curriculum. The curriculum has two pre-clinical and four clinical years, one of which is an internship year taken outside the university setting as a pre-requisite for full medical licensure. During the clinical attachment, trainees complete one to five months of supervised rotations in major departments, including but not limited to; surgery and traumatology, obstetrics and gynecology, internal medicine, and pediatrics. In principle these medical trainees provide the first contact with trauma patients. As such, they complete patients’ admission, initiate emergency resuscitation, discuss case management amongst themselves (horizontal consultation) before consulting with senior faculty, and may discharge or coordinate patients’ referrals. In the face of scarce human resources, it is therefore considered that incorporating these trainees into trauma teams under guidance with “pre-and in-service” skills training, could formulate consistently structured-priority-driven injury care. Research in Sweden shows that interns are underutilized powerful catalyzers for medical learning, and for taking responsibility in patients care pathways ^17^.

There have been efforts to incorporate an advanced trauma life support course (ATLS) of the American College of Surgeons ^18^ in Ugandan medical curricula, however the content remains theoretical without being conceptualized to suit the country’s resource limited settings. In addition, ATLS in its patented form is arguably expensive for medical trainees in LMICs. To-date, Uganda does not have any certification center for this program, requiring one to travel overseas to attain this valuable training. There were further efforts to scale out an alternative level III targeted primary trauma courses (PTC) in Uganda ^19^, when it demonstrated its potential in skill improvement in India ^20^, but the course failed due to lack of international accreditation and recognition for global job markets in addition to its minimal reported impact on mortality reduction ^16^. The more applicable, internationally recognized rural trauma team development course (RTTDC)^21^ of the American College of Surgeons ^22^ has demonstrated its potential in minimizing morbidity and mortality amongst a general trauma population ^23^, but has been evaluated in middle and high-income countries ^24^.

According to the Uganda police annual traffic report (2017), 78% of fatal road traffic injuries (RTIs) occur in rural regions where doctors are scarce and where police ambulances are the most accessible ^25^. The Ugandan ministry of health and medical schools purposively deploy medical trainees in such rural regional referral hospitals (Level III trauma centers) partly to address the human resource needs, and to accelerate the trainees’ “hands-on” experience. Therefore, building capacity for rural trauma training to the traffic law enforcement professionals who undertake the role of pre-hospital emergency evacuation of injured patients from rural regions and to medical trainees who provide the first hospital-based emergency care would positively impact the consequences of injuries in such environment ^23^. However, the inception, implementation, and outcome impact of rural trauma team development training in such settings is poorly documented. To the best of the authors’ knowledge, Ugandan studies which have evaluated the applicability and impact of such an intervention are scarce. In this study, undergraduate medical trainees and traffic police are considered potential human resources for immediate trauma care.

### Study Objectives

#### Main objective

The main objective of this study is to evaluate the feasibility of rural trauma team development training amongst medical trainees and road traffic law enforcement professionals, using the rural trauma team development course (RTTDC)^21^ of the American College of Surgeons ^18^ as a starting point. This research is nested in a broader ongoing trial on the use of allied health and law enforcement trauma registries for motorcycle related injuries in Uganda (Pan African Clinical Trial Registry No. PACTR202308851460352). The target course participants were chosen for this intervention because they are often the first responders in trauma care in Uganda’s resource constrained settings.

#### Specific objectives

i. To compare the pre-and post-RTTDC training mean scores amongst medical trainees.
ii. To determine the relevance and acceptability to promulgation of RTTDC amongst medical trainees and traffic law enforcement professionals in Ugandan rural level III trauma centers.

##### Hypotheses

The null hypothesis of this research protocol is that there is no difference in pre-and post-interventional training mean scores of research participants.

## Methods

### Study design

This will be a multi-centre interrupted time-series of interventional trainings in a prospective cohort of medical trainees and traffic law enforcement professionals during 28^th^ September 2019-28^th^ November 2023. This protocol is developed in accordance with the (STROCSS, 2021) guidelines for strengthening the reporting of cohort, cross-sectional and case-control studies in surgery ^26^; and in line with guidelines for reporting evidence-based practice educational interventions and teaching (GREET)^27^.

### Source of data and study settings

This study will be conducted at three regional referral and teaching hospitals in Uganda, including Kiryandongo, Jinja and Hoima. These facilities have similar characteristics in the sense that they all serve as internship and residency sites for graduating general doctors, surgery residents and as teaching sites for the Kampala International University Medical School. In cooperation with local surgeon consultants, each of these tertiary hospitals offers specialized services including emergency surgery for injured patients such as those with neuro, orthopedic, and general trauma. The key staff in the surgery and trauma departments include at least a specialist surgeon, a master’s resident, intern doctor/nurse and support staff who work with other departments such as occupational therapy, physiotherapy, and rehabilitation to offer 24-hour multi-disciplinary services to the injured patients. In addition, each of these facilities receive undergraduate medical trainees including those studying for Bachelor of Medicine and Bachelor of Surgery, Bachelor of Nursing Sciences, and Bachelor of Clinical Medicine degrees here being collectively referred to as medical trainees. The trainees rotate on a three-months basis in major clinical disciplines such as surgery amongst others. Further, each of these hospitals are located directly opposite the regional district police headquarters which offers complementary services such as legal guidance on traffic crashes, third party compensation claims and issuance of “police form 3A” for the examination of injured persons. Patients sustaining either intentional or unintentional injuries are immediately encouraged to report to the police for investigations and often police attend to such patients as first-responders at the crash scenes.

### Scope of the study training

Rural trauma team development course was developed by the committee on trauma of the American College of Surgeons^21^ to help rural health facilities improve on the quality of injury care and trauma outcome. It was designed on the basis that in most situations, rural facilities can form an organised efficient core response team consisting of at least 3 members for example doctor (team leader), allied health professional (such as a medical intern or nurse), paramedic (rescue traffic police in Ugandan settings) to address the healthcare needs of an injured patient. This face-to-face training will entail a brief introduction to rural trauma systems, pre-test questionnaire regarding pre-existing knowledge about the subject matter, essentials of pre-hospital communication to receiving hospitals and safe transportation of a trauma patient (targeted for traffic police who often undertake this role in the Ugandan context). Further, we shall organise participants in teams of five with specific roles such as team leader or member, identify local resources and limitations, and demonstrate resuscitation of trauma patients through simulated case scenarios with locally available: resuscitation equipment, mannikins, radiographs, PowerPoint projections and videos. The efforts throughout the training will endeavor to demonstrate to participants a sequential approach to trauma, i.e., primary survey, decision to transfer, secondary survey and process improvement by use of clinical case scenarios.

Subsequently, teams will be tasked to manage trauma case scenarios during which they will be observed for the following skills. First, to perform a primary survey to demonstrate assessment and interventions for airway compromise with emphasis on skills for manual airway maneuvers. Secondly, to demonstrate teamwork, leadership, and communication skills in making timely decision to consult senior faculty or transfer. Thirdly to execute secondary survey and demonstrate assessment for respiratory distress, quality of ventilation, oxygen administration, chest-tube and needle thoracotomy, 3-way occlusive dressing, detection of shock, control of external hemorrhage with compression or tourniquet, fracture splintage and spine immobilization for safe transfer, detection of deteriorating level of consciousness and clinical signs of raised intracranial pressure. Lastly, to provide peer review, audit team performance, and suggest areas for improvement to increase patients’ safety with particular attention on how the team’s approaches would differ in special trauma populations such as pediatrics, elderly, and pregnant women.

Participants will be given expert feedback on their team performance regarding case scenarios. First, based on the assigned skilled team member roles as per the RTTDC 4^th^ edition ^21^. Secondly, based on the trauma non-technical skills scale (T-NOTECHS) ^28^. The (T-NOTECHS) scale rates team performance based on five domains including leadership; cooperation and resource management; communication and interaction; assessment and decision-making, and situation awareness and coping with stress. Each domain will be evaluated on a Likert scale of 1-5 where 5 represents an ideal flawless team performance whereas 1 indicates performance that did not demonstrate teamwork behaviour as described by Steinemann et al ^28^. This tool has been validated and found to be reliable for evaluation and measurement of non-technical skills during training of trauma teams ^29^. In return, participants will give their immediate feedback on the training through a structured post-course evaluation form and complete the post-training trauma based MCQs at 3-months follow-up. The training and assessments will be conducted in English which is the official language of instruction in Uganda.

### Training procedure, team construct and course implementation

On average 12 (6-24) training sessions will be conducted per site between 28^th^ of September 2019 and 28^th^ of November 2023. The trainings will follow a 3-months cycle, corresponding to the duration of surgical internship clinical rotations and deployment by the Ugandan ministry of health and medical schools. Further, the effects of the training are presumed to “decay” after this period and the authors project to realize the targeted sample size based on the 3-months interval. However, permission will be sought from the ethical committee to extend the study in case the target sample is not obtained during the approved duration of the study. The mode of delivery and content of the RTTDC training will not be modified. The training will be advertised by hospital administrators and class representatives and eligible participants will be recruited based on first come first serve. The training dates will be selected to suit the designated days for clinical grand rounds and/ or continuous professional development sessions so as not to interfere with the university and hospital daily schedules. The investigators will use student unique numbers from university hospital administrators’ database to randomly assign recruited participants into teams and to ensure that each participant is recruited once. The participating law enforcement professionals who will show up for the training will pick ballot papers randomly to be assigned to the already created medical trainee teams. The chief trainer will be a chief surgeon (HL) who attended the RTTDC instructor course. The surgeon will coach two co-trainers who are master of surgery residents undertaking a traumatology rotation, at the participating hospitals to make a total of three trainers. Since the program is aimed at capacity building, and residents directly supervise interns in Uganda, the research and ethics board approved training of residents as opposed to consultants not to over strain the skeleton specialized staff at the study sites. Interns and residents contribute to 75% of the health workforce in public hospitals in Uganda ^8^. The program will be designed for two days, targeting 30 participants with a trainer: trainee ratio of 1:10, which is the maximum permissible for each training session according to the committee on trauma of the American college of Surgeons ^21^. For each team task, participants will be grouped into teams of 5 to role play a surgeon, surgeon assistant/intern, an anesthesiologist/anesthetic assistant, an allied health professional (emergency room nurse/laboratory/imaging personnel), and a rescue police (representing paramedics in Uganda’s settings).

### Course outcomes

At the end of the training, participants are anticipated to be able to describe components of their current local trauma system (trauma capacity), describe barriers to effective trauma care in their settings, suggest areas for performance improvement, demonstrate concepts of primary survey and secondary survey, know when to make a timely decision to consult or transfer (referral) for definitive care, and understand the link between lower hospital facilities and regional trauma centres. These goals have been set apriority in accordance with the committee on trauma ^21^. The participants will complete a paper-based pre-training trauma related MCQs before and at 3 months after the training. The method of data collection will remain the same with a target data completion rate of 100% for every series. The pre-post training MCQs will be marked by two independent consultant surgeons who are not part of the research team. The marking team will be blinded by what is pre-or-post training. Since the answers and marking guide are standardized, we anticipate an inter-rater agreement of 100%.

### Study population

We shall target nursing students and medical students/interns who are often the first contacts to trauma patients in hospital settings and road traffic law enforcement professionals who are often involved in transportation of such patients to hospitals. The accessible population will be those medical trainees/interns completing the surgery rotation during the study period and police officials assigned to complete the training by virtue of their roles.

### Sample size estimation

According to the respective hospital administrators, a total of 1500 medical trainees had been received at the three participating hospitals during the academic year 2018/2019 which preceded the study. Using the hypergeometric formula in equation 1 for determining sample size in smaller populations,

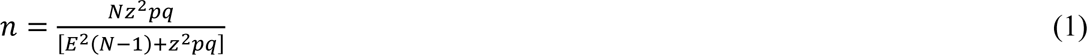

Where:

N (population size) =1500

Z=1.96 assuming 95% confidence interval

P and Q are population proportions that will approve or disapprove the promulgation of rural trauma team development training, each assumed to be 50% since these proportions are unknown E = value of accuracy of the sample proportions (alpha= 0.05) at 95% confidence interval n (sample size) =307. We shall add 41% to increase internal validity for the 4-year period to cater for the projected 10.25% annual loss to follow-up and non-response, thus the final minimum sample size for medical trainees will be 433.

### Sampling methods

For proportionate representation, the proportion “p” will be computed as shown in equation 2, depending on the number of medical trainees each hospital received during the year that preceded the study.

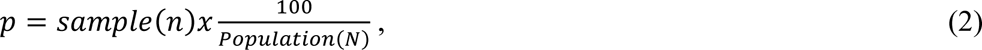

This means that approximately 26.7% of each hospital’s annual trainees intake will be recruited, resulting in 321 participants from Kiryandongo, 65 from Jinja and 47 from Hoima regional referral hospitals.

For the case of law enforcement professionals, the selection will be purposive where the regional police in-charge of traffic will be asked to nominate 22 participants for each region totaling to 66, targeting police stations on main highways leading to the respective municipalities where the study sites are located. Those who consent to participate will then be randomly assigned to join the medical trainee teams.

### Inclusion criteria

We shall include participants who meet the following criteria: road-traffic police officer, medical intern doctor/nurse, medical student enrolled in Bachelor of medicine and Bachelor of Surgery in clinical years (3^rd^ or 5^th^) who completed or currently is completing the surgical/traumatology clinical rotation, allied-health professional trainee enrolled in Bachelor of Nursing Sciences or Bachelor of Clinical Medicine degree program and is involved in the care of the injured patients or attached to the accident and emergency department during the study period.

### Exclusion criteria

We shall exclude participants who know would not complete all modules of the course for foreseeable reasons such as students having an exam on one of the training days or police officers on duty on the day of training as this would have an impact on the overall course evaluation. Such individuals will be provided with alternative dates (if available).

### Quality control, study variables, data collection tools and their validity

A standardized 4^th^ edition of RTTDC will be delivered without modification in which 20 item pre- and post-course multiple choice questionnaire (MCQ) will be administered to participants based on the 4^th^ edition of RTTDC instructor manual ^21^. Video recordings will be used to capture skill-oriented case scenarios. Desirability of this training and its content will be determined using a 5-point-Likert scale (5=strongly agree, 4=agree, 3=neutral, 2=disagree, 1=strongly disagree) on the 16 topics whose reliability were validated earlier ^30^. The relevancy of quality improvement, case scenarios and communication course modules to the study subjects will be rated on a 3-point scale based on their subjective impression as (3=very relevant, 2=relevant, 1=not relevant), in conformity with previous researchers ^30^. Post course feedback on team performance will be provided in accordance with the standard RTTDC expected trauma technical team member assigned skills and the validated (T-NOTECHS) ^29^. Three trainers will consistently deliver each session of the training throughout the study period to minimise inter-operator variation. Team members will maintain their team-mates on both training days. Two offsite independent surgeon consultants will evaluate the MCQs while blinded of which is pre-or post-training test. The marked items will be returned to an offsite study administrator with an MCQ marking guide who will double check the awarded marks prior to data entry and disqualify any participant with wrongly marked MCQs from inclusion in the final analyses. The data will be collected in hard copies and later transferred to the Research Electronic Data Capture (REDCap) tools hosted by the University of Turku. REDCap is a secure, web-based software platform which provides an intuitive interface for validated data capture and audit trails for tracking data manipulation ^31^. Further, the software enables seamless data export and download procedures that are compatible with commonly used statistical packages, while allowing for data integration and interoperability with external sources ^32^.

### Statistical Analysis

The data will be exported from REDCap to Stata version 15.0 for analysis (StataCorp. 2017. Stata Statistical Software: Release 15. College Station, TX: StataCorp LLC).

We shall test the data for normalcy using the Shapiro-Francia test and normal histogram curves at 95% CI, regarding p<0.05 as statistically significant.

If the data are normally distributed, the difference in pre-and post-training mean scores will be compared using one way analysis of variance (ANOVA) test otherwise non-parametric Friedman test will be used to compare the scores as a measure of knowledge retention, assuming a null hypothesis of no difference.

Further, we shall perform Wilcoxon signed rank non-parametric test to examine the difference in the effect the training has on MCQ scores of the various cadres of participants, aggregated as (3^rd^ year, 5^th^ year, interns). The median scores for various groups and their corresponding interquartile range will be reported.

The statistical difference in rank sums regarding desirability and relevance of various course themes will be compared between participating cadres using a non-parametric Kruskal Wallis (H) test.

Lastly, time series regression models will be used to test for autocorrelation trends in MCQ performance over the study period for the various medical trainee groups.

All the analyses will be performed using Stata 15.0. (StataCorp. 2017. *Stata Statistical Software: Release 15.* College Station, TX: StataCorp LLC); at 95% confidence interval, regarding a 2-tailed p-value <0.05 as statistically significant.

### Statement of ethics approval and consent to participate

The study will be executed in accordance with the US National Institute of Health guidelines for research involving human subjects as participants; and all methods will be performed in accordance with comparable ethical standards as laid down in the Declaration of Helsinki ^33^ and its later amendments. Prior to recruiting participants, ethical clearance was obtained from Mbarara University of Science and Technology (Ref: MUREC 1/7; 05/5-19) and final ethical approval was granted from the Uganda National Council for Science and Technology, the authority which regulates research in Uganda (Ref. No. SS 5082). It will be pre-requisite for each eligible participant to endorse a pre-designed informed consent form with their signatures in the presence of principal investigator or designated trainers prior to participation in the study. The official consent form for Mbarara University of Science and Technology will be adopted for this purpose. Participation will be free and voluntary, and participants will have the right to withdraw from the study at their wish. Video recordings will be destroyed 4 months after expert feedback on team performance and retrieval of main content themes for each cycle of training.

### Participant recruitment workflow

The potentially eligible participants will be registered, briefed, and educated about the study two weeks prior to the training; and will be availed with links to the educational materials. Consented participants will be recruited for this two-day event. On the agreed first day of the training, interns and medical trainees will complete pre-training MCQs during the first 40 minutes in “an exam-like” supervise environment, followed by assignment into random groups. The training will be divided into four modules i.e., (i) introduction to rural trauma systems, primary and secondary survey, (ii) Case scenarios, (iii) trauma communication and (iv) patient safety and process improvement. Each module will be allocated two hours to make a total of eight hours of “continuing medical education” which is the maximum allowable by the committee on trauma of American College of Surgeons^21^. The first half of the course modules will be delivered during the opening day and the remaining half on the second day of the training.

The training for all teams will take place between 9:00am and 15:00pm and the venue will be the designated spacious multimedia surgical simulation room located at each of the training hospitals. There will be coffee breaks of 45 minutes between each 2-hour session on each day of the training. The PI will cater for the coffee budget to enhance attendance and time efficiency. The participants will maintain their teams throughout training days. Study participants will be given overall team performance feedback after the last module and each study subject will be asked to complete a 15-munutes course evaluation form regarding acceptability and relevance of the training at the end of the second day. Subsequently, trainee participants will be followed up after 3 months (90 days) from the first day of the training to complete a 40-minutes post training trauma based MCQs in “an exam-like” supervised environment as shown in Figure 1.

**Figure 1:**
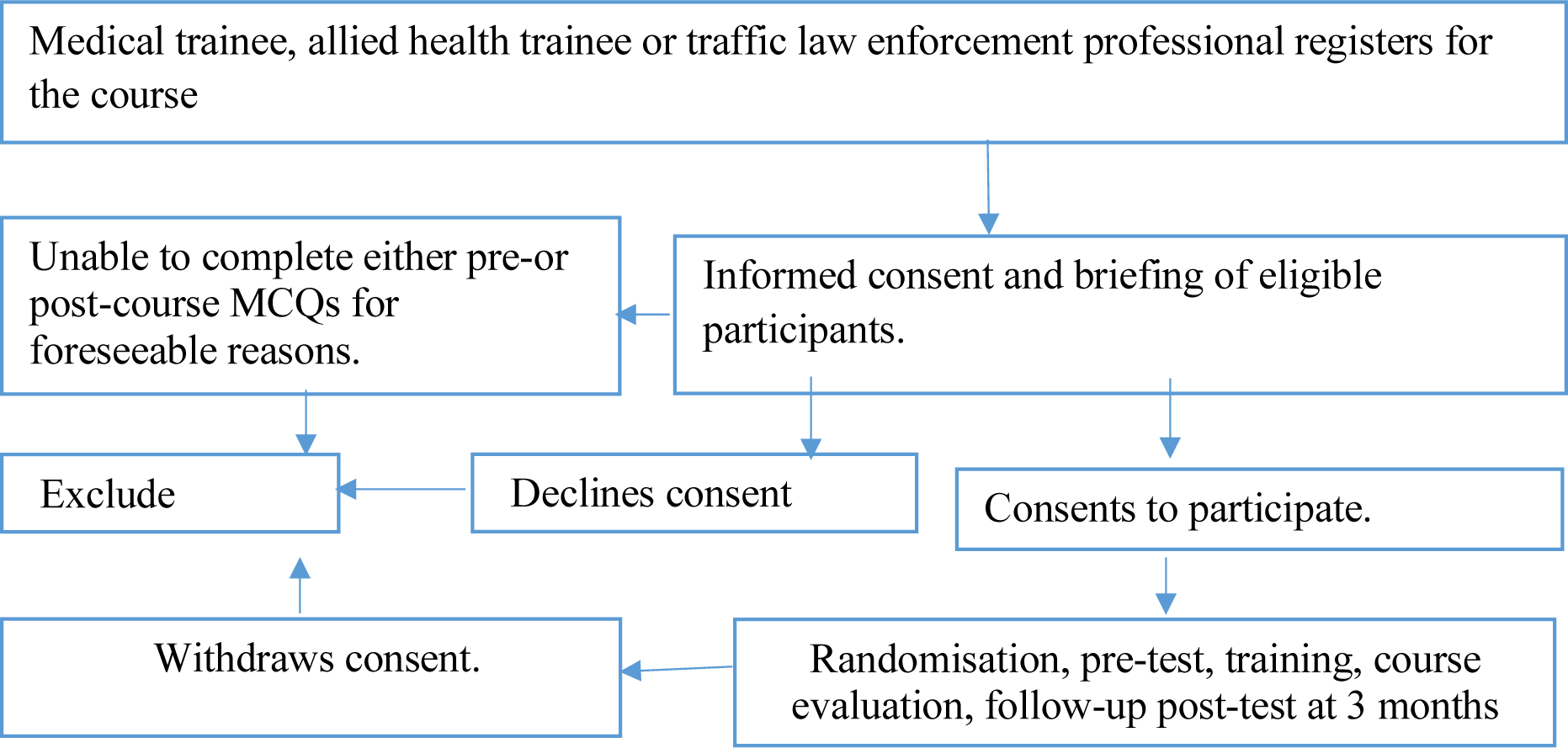
Flow diagram showing recruitment process of study participants.

## Discussion

Compelling evidence from high income countries suggest that rural trauma team development course training could improve the efficiency of care and clinical outcomes of patients who sustain trauma ^34,35^. In their results, scientists demonstrated that these benefits of rural trauma team training could arise through improved clinicians’ knowledge, and team organization in terms of leadership, communication and quick decision making; subsequently shortening the time to execute treatment or transfer the injured patient to higher level trauma centers ^22,23,36^. However, what is defined as “rural” in USA settings is not depictive of LMICs. For instance, whereas the decisions for pre-hospital care and emergency evacuation of injured patients are made by emergency physicians and paramedics, using air-ambulances in high income countries ^35^, these tasks are often executed by allied health, medical trainees and traffic law enforcement professionals often using public transport in Uganda ^37,38^. This timely study will examine the conceptualization of rural trauma team development through providing training capacity to traffic law enforcement and trainee medical professionals who are viewed as potential sustainable human resource for injury care in low-resource settings. Our findings on acceptability and applicability of the proposed training could potentially inform the design of future rural trauma teams in real-world clinical settings of LMICs. Further, when our study findings become available, the results could guide the design of traffic law enforcement and medical curricula in Uganda and similar low resourced settings.

### Study strengths and limitations

This multi-center study is among the few long-term studies to evaluate the effect of rural trauma team development training amongst rural trainee medical participants and traffic law enforcement professionals in typical low-income settings. However, there will be some limitations in this study. First, our trauma team simulation with traffic police and students in medical training could differ from the physician-led emergency medical teams in affluent Western countries, consequently limiting the generalizability of our results to LMICs. Secondly, regarding training for police, we hope to emphasize the key communication aspects of evacuation and prehospital safe transportation of injured patients to hospitals. As such, we shall not assess the trauma knowledge based MCQ for law enforcement participants on assumption that they have no pre-existing clinical exposure but information on the applicability of the gained knowledge to their work environment will be obtained. Furthermore, medical trainee participants’ knowledge could change with time irrespective of the intervention, but we hope that having multiple interrupted time series of intervention sessions at multiple study sites and using time series autoregression models will overcome this obstacle in addition to limiting confounding due to between group differences^39^. Lastly, the mean difference in pre-post training MCQs for medical trainees might only depict knowledge improvement which does not necessarily transform to clinical skills that impact patient outcomes. However, the authors plan to transfer this classroom knowledge into clinical practice in a cluster randomized controlled trial to determine whether the apparent knowledge changes during the training are not due to other interventions, impact on clinical time efficiency, and patient outcomes in our constrained resource context.

## Data accessibility statement

All the de-identified data sets arising from this study protocol will be made publicly available as online supplemental material through a permanent weblink to a repository that will be provided by peer reviewed journal. The data collection instruments are available through the corresponding author.

## Author contribution

HL Principal investigator; HL, MLW Conceptualization; HL Data curation, Investigation, Methodology, Project administration, Resources; HL, MM Formal analysis, Software, Visualization; HL Writing-original draft. RS, PK, JPP, MLW Validation, Writing-review, and editing; JPP, MLW Supervision, Funding acquisition. All authors read and approved the final manuscript for submission.

## Conflict of interests disclosure

All authors report no competing interests to declare. The content of this paper is the sole responsibility of the authors and do not represent any official views of their institutional affiliations. The training simulation for participants in this study is not for any formal academic accreditation or institutional awards, neither does it represent any official views of the American College of Surgeons. The research and ethics committee approved this educational activity with a maximum of 8 CPD points.

## Sources of funding

JPP is supported by the Academy of Finland (Grant No. 17379) and the Maire Taponen Foundation. The study sponsors did not have any role in the design, writing or decision to submit the study protocol for publication.

## Research registration unique identifying number

Since the country of primary recruitment where the study was approved does not have a publicly available electronic register, this protocol has been retrospectively registered with research registry UIN researchregistry9490 hyperlink https://www.researchregistry.com/browse-the-registry#home/

## Study updates

Actively recruiting, first participant recruited 28 September 2019, last participant anticipated by 28 November 2023

## Guarantors

HL and MLW.

## Data Availability

All data produced in the present work are contained in the manuscript

https://www.researchregistry.com/browse-the-registry#home/

## Acknowledgements

The authors are grateful to the staff of Mbarara University of Science and Technology Research Ethics Committee for providing constructive ethical guidance and academic scrutiny during the early phase of this work.

## Provenance and peer review

Provenance and peer review not commissioned, awaiting external peer review in International Journal of Surgery Protocols.

## Notes

### Competing Interest Statement

The authors have declared no competing interest.

### Clinical Trial

PACTR202308851460352

### Clinical Protocols

https://www.researchregistry.com/browse-the-registry#home/

### Funding Statement

This study did not receive any funding

### Author Declarations

Research and Ethics Committees of Mbarara University of Science and Technology (Ref: MUREC 1/7, 05/05-19) and Uganda National Council for Science and Technology (Ref: SS 5082) gave ethical approval for this work

